# Methods for estimating maternal, newborn, and child health and nutrition effective coverage cascades from household and health facility surveys

**DOI:** 10.1101/2024.12.20.24319361

**Authors:** Melinda K. Munos, Ashley Sheffel, Emily Carter, Jamie Perin, IMPROVE Coverage Group

## Abstract

**Background:** Effective coverage cascades have been proposed to understand to what extent populations are able to benefit from interventions to address their health needs. Theoretical effective coverage cascades have been developed for reproductive, maternal, newborn, child, and adolescent health and nutrition (RMNCAH&N), but there is no consensus regarding the methods to estimate effective coverage cascades. We operationalized the proposed effective coverage cascades for selected RMNCAH&N services; this paper presents the overall methods, challenges, and lessons learned.

**Methods:** We used data from Demographic and Health Surveys, Multiple Indicator Cluster Surveys, Service Provision Assessments, and the Service Availability and Readiness Assessment to estimate effective coverage cascades in seven low- and middle- income countries for the following service areas: antenatal care, care for small and/or sick newborns, postnatal care, sick child care, and maternal and child nutrition. We developed operational definitions for each of the seven steps of the effective coverage cascade and developed readiness, and, where data allowed, process quality indices for each service area. Readiness- and process quality-adjusted coverage were estimated using ecological linking by stratum. We propose approaches for dealing with multiple observations per facility; multiple care-seeking episodes; and empty strata, as well as a jackknife approach to estimate the standard errors for readiness- and process quality-adjusted coverage.

**Results:** We were able to estimate effective coverage cascades through intervention coverage (step 4) for postnatal care and through process quality-adjusted coverage (step 5) for antenatal care, sick child care, and maternal and child nutrition. For small and/or sick newborn care, we did not have an appropriate denominator or measure of service contact coverage and had to modify the cascade significantly. Data gaps were the largest barrier to the estimation of effective coverage cascades for RMNCAH&N. Other challenges included accounting for community- and home-based interventions, determining whether the cascade should be nested, and interpreting the cascade.

**Conclusions:** To make effective coverage cascades feasible for routine use, clear guidance is needed on cascade methods and definitions, accounting for the full spectrum of RMNCAH&N interventions, and developing our understanding of how coverage cascades can be used by stakeholders to improve health systems and programs.

## Background

Since 2000, the world has made substantial progress in improving the health of mothers, babies, and children: under-five mortality has dropped from 93 per 1000 live births to 38 per 1000 globally [1], the proportion of children under five years with chronic malnutrition (stunting) has declined from 33% to 22% [2], and the maternal mortality ratio has declined from 339 to 223 deaths per 100,000 live births [3]. Despite these improvements, levels of child and maternal mortality, morbidity, and malnutrition remain high in many settings, particularly in lower-resource settings and among marginalized populations with limited access to health services [4]. To improve health status, it is essential to understand to what extent populations are able to access and benefit from interventions and services to address their health needs.

Intervention coverage, or the proportion of individuals in need of an intervention who receive that intervention, has been extensively used to assess who is being reached by interventions, but it has a few shortcomings. In some cases, measures of service contact coverage – the proportion of a population in need who access a service such as antenatal care, rather than a specific intervention – have been prioritized. However, service contact coverage does not provide information on which interventions were actually received, and neither intervention coverage nor service contact coverage account for the quality of care received. As a result, effective coverage (EC) has been proposed as an approach to understand the health gains accruing to populations from interventions and services, and the bottlenecks to achieving those health gains [5–7].

The EC cascade was first proposed by Tanahashi in 1978 [8] and further developed and adapted for reproductive, maternal, newborn, and child, and adolescent health and nutrition (RMNCAH&N) by Amouzou and colleagues in 2019 [5]. The cascade follows a population in need of a health service or intervention, identifying bottlenecks and missed opportunities that impede the target population from fully benefitting from the service or intervention. It includes (1) the proportion of the target population that (2) visits a health service, (3) which is “ready” to provide the interventions needed, (4) receives the necessary interventions, (5) receives the interventions according to quality standards, (6) adheres to the interventions, and (7) has a positive outcome or health gain [5]. In 2019, the World Health Organization (WHO) and the United Nations Children’s Fund (UNICEF) convened a consultation – the Effective Coverage Think Tank. The resulting publication by Marsh et al. proposed consensus definitions for EC cascades and their components, as well as example or theoretical EC cascades for certain RMNCAH&N service areas [6].

Studies have attempted to estimate EC using various definitions and methods [9], but there has been limited experience applying the EC Think Tank definitions using empirical data, and there are as of yet no consensus recommendations on the specific methods to estimate these cascades. Building on the work of the EC Think Tank, we aimed to operationalize the Think Tank and Amouzou cascade definitions for selected RMNCAH&N services using existing household and health facility survey data to examine the feasibility and methods necessary to estimate EC cascades. The service area-specific cascades will be published separately. This paper describes the overall methods that were used for estimating these EC cascades, and discusses the challenges we encountered and recommendations for future EC research.

### Terminology and Definitions

We used the coverage cascades proposed by Amouzou et al. and by the EC Think Tank (Marsh et al.) as our generic conceptual cascade [5,6]. **Table 1** provides the conceptual definitions for each element of the cascade as adapted from Amouzou et al and Marsh et al, as well as the operational definitions used in our analyses. For each service area, we then defined a conceptual cascade based on this generic cascade and data availability.

**Table 1.** Conceptual and operational definitions of the effective coverage cascade components.

| Cascade component (Marsh) | Conceptual definition (slightly adapted from Amouzou) | Operational definition | Data Source used | Notes |
| --- | --- | --- | --- | --- |
| Target population | All who need a service/ intervention | No change | Household Survey (DHS/ MICS) | Need may be defined based on various factors including demographic status (age, sex, pregnancy) or health status (nutritional status, various diagnoses). |
| Service contact coverage | Proportion of the target population who visit a health service | Proportion of the target population who visit a health facility or **qualified provider** | Household Survey (DHS/ MICS) | Qualified provider should be defined for each country and service area, and may include public sector, non-public sector, facility, and non-facility (e.g., community health workers) providers, depending on country policies and the particular service area or intervention. |
| Input-adjusted coverage | Proportion of the target population who visit a health facility or provider that is 'ready' (i.e., all necessary inputs are available) to deliver the required intervention(s) or services | Each qualified provider or health facility is given a mean readiness score. Input-adjusted coverage is the proportion of the target population who visited a health facility or qualified provider, scaled by their mean readiness score. | Linked Household Survey (DHS/ MICS) and Health Facility Survey (SPA/ SARA) | This is sometimes called "readiness-adjusted coverage". Facility or provider readiness is calculated on a scale of [0,1] with 1 being perfect readiness and 0 being no readiness. Readiness is defined for each intervention/service and setting, but typically accounts for infrastructure, human resources, drugs, diagnostics, commodities, other supplies, and guidelines needed to deliver the service or intervention according to standards. |
| Intervention coverage | Proportion of the target population who received the needed health intervention(s) or services | No change | Household Survey (DHS/ MICS) |  |
| Process quality-adjusted coverage | Proportion of the target population receiving the intervention(s) or service according to recommended standards (provider adherence to standards). | Proportion of the target population who visited a health facility or qualified provider, scaled by the mean quality score. | Linked Household Survey (DHS/ MICS) and Health Facility Survey (SPA/ SARA) | Facility or provider quality is calculated on a scale of [0,1] with 1 being perfect quality and 0 being no quality. Quality of care is defined for each intervention/service and setting, but accounts for whether the patient received the intervention or service according to clinical standards (including, e.g., appropriate history taking, assessment, counselling, use of diagnostics and treatment if appropriate) according to standards. |
| User adherence-adjusted coverage | Proportion of the target population receiving the intervention(s) or service according to recommended standards and adhering to the treatment guidelines. | Not estimated | Not estimated | Adherence is not relevant for all interventions; in cases where the intervention is entirely delivered by the provider and there is no home care, behavior change, or continuing treatment needed, this component can be omitted. This is also difficult to estimate for a package of services. |
| Outcome-adjusted coverage | Proportion of the target population experiencing the health gains from the service. | Not estimated | Not estimated | Measured RMNCH&N outcomes are broad and difficult to attribute to an intervention or service area. |

A few notes on terminology: in line with Marsh et al., we define “effective coverage” as outcome-adjusted coverage, i.e. the “proportion of the population in need of a service that received a positive health outcome from that service” [6]. We use the term “effective coverage cascade” (or EC cascade) to refer to the set of steps leading to outcome-adjusted coverage as described in **Table 1**, excluding those for which no data were available (these cascades have also been referred to as “coverage cascades” or “health service coverage cascades” [5,6]). We use “readiness” to refer to input or structural quality [10], “process quality” to refer to the provision and experience of care [11], and “quality of care” to refer to readiness and process quality as a whole.

### Selection of data sources, service areas, and countries

#### Data sources

The data sources for these analyses were household and health facility surveys. For the household surveys, we used the Demographic and Health Surveys (DHS; https://dhsprogram.com/) [12], and Multiple Indicator Cluster Surveys (MICS; https://mics.unicef.org/) [13] datasets (**Table 2**) to define the target population, calculate care-seeking and intervention coverage, and identify sources of care, as described below.

**Table 2.**
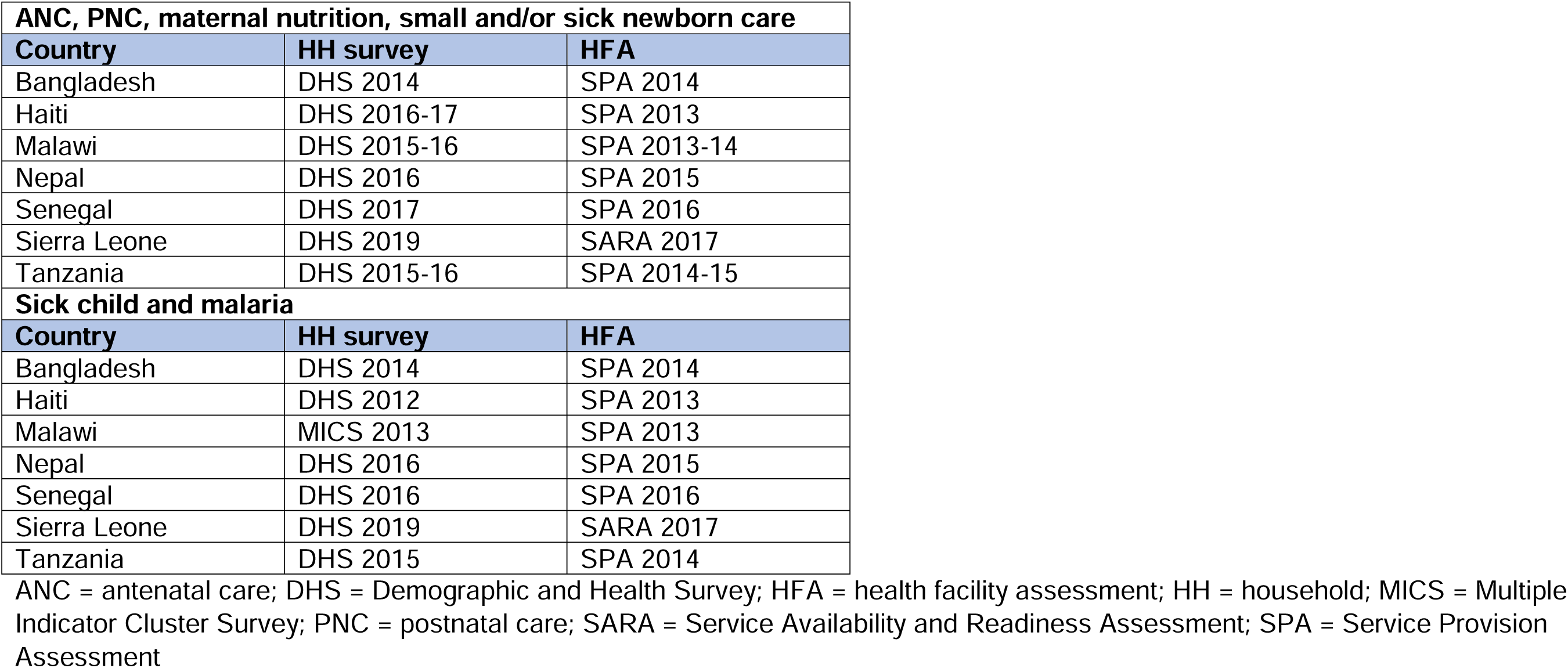
Data sources.

More information on the DHS and MICS is available on their websites and in the survey reports [12,13], but briefly, the DHS is a stratified multi-stage household survey that collects data on demographic and health data for households and individuals. The sample is typically stratified by sub-national administrative area and by urban/rural area of residence. Within each stratum, clusters are sampled with probability proportional to population size, and systematic random sampling is used to select households in each sampled cluster. The woman’s questionnaire identifies women with a recent live birth and with children aged less than five years, and asks questions about women’s antenatal, childbirth, and postnatal care, as well as the health services and interventions received by their children. DHS provides population-based estimates of service contact coverage and intervention coverage at national and sub-national levels. The MICS is similar to the DHS, but with a slightly different questionnaire structure and set of questions, and a shorter reference period for maternal indicators. Notably, the MICS does not collect data on of the type of facility where women received antenatal care (ANC), so we did not use MICS data to calculate readiness- or quality-adjusted coverage for ANC.

We used Service Provision Assessment (SPA; https://dhsprogram.com/Methodology/Survey-Types/SPA.cfm) and Service Availability and Readiness Assessment (SARA; https://www.who.int/data/data-collection-tools/service-availability-and-readiness-assessment-(sara)) data to estimate facility readiness and process quality [14,15] (**Table 2**). More information on the SPA and SARA (which has been replaced by the Harmonized Health Facility Assessment (HHFA)) is available on their websites and in the survey reports [14,16]. Briefly, the SPA is a survey of health facilities drawn from a comprehensive list of health facilities in the country. The list is typically stratified by health facility type/level and managing authority (i.e. public, private, faith-based, etc.) and facilities are randomly selected. At each sampled facility, a sample of health workers are interviewed about their training and supervision, and a sample of family planning, ANC, and sick child consultations are observed and exit interviews are conducted. In addition, the SPA includes a health facility inventory that collects data on service availability and availability of equipment, drugs, and commodities. The SARA is similar to the SPA except that it includes only a facility inventory. Facility readiness data were obtained from the SPA and SARA facility inventories, as well as the SPA health worker interview data. Process quality data were obtained from the SPA ANC and sick child observations and exit interviews. Where possible, we preferentially used SPA data rather than SARA because SPA includes observations of ANC and sick child consultations, allowing us to estimate process quality-adjusted coverage for these service areas, and includes more detailed data on human resources than SARA, including individual-level data on staff training.

We considered using routine health information system (RHIS) data but ultimately decided to focus on publicly available data that are standardized across countries for this initial cross-country analysis. However, exploring the use of RHIS data to estimate EC cascades for national quality planning is an important next step that would allow for more frequent analyses of EC cascades in more countries. A small number of studies have explored the use of RHIS data for effective coverage [17].

#### Service areas

We estimated EC cascades for ANC, postnatal care (PNC), care for small and/or sick newborns, sick child care, and maternal and child nutrition. These service areas were selected to include a range of RMNCAH&N life stages, preventive and curative care, and different levels of data availability.

#### Countries

We included low- and middle-income countries (LMICs) with recent, publicly available, national household and health facility datasets that could be linked to each other (as described below). We identified all countries with a DHS or MICS survey in the five-year period covering 2013 to 2018 (when we started these analyses) and a publicly available health facility survey conducted in the same year as, or in the two years prior to the household survey. The two-year restriction was necessary because we linked care-seeking data from the household survey to readiness and process quality estimates from the facility survey (**Table 1**). The household survey care-seeking data have reference periods ranging from two weeks to two years before the survey, depending on the target population. To appropriately link the two datasets, the health facility data needed to be collected during or close to the relevant reference period for the household survey dataset.

Although many countries had a national household survey within this time period, only eight countries also conducted a national facility survey with publicly available data from 2011 to 2018. Two countries were excluded because there was no household survey conducted in the 2 years prior to the HFA (Afghanistan and Haiti; we later included Haiti as described below). One country was excluded because the data were not yet publicly available in 2018, when we began our analyses (Democratic Republic of Congo). One country (Haiti) had conducted a SPA three years prior to a household survey, but we included it to increase the geographical representativeness of the analysis. Several countries conducted a SARA during this time period, but these datasets are not publicly available. Because we aimed to include countries that were geographically diverse, we also sought out a West African country that could provide access to de-identified SARA data that could be used in these analyses. We received permission from the Sierra Leone Ministry of Health and from the World Health Organization (WHO) to use the 2017 Sierra Leone SARA, which we linked to the 2019 Sierra Leone DHS.

### Measurement approach for estimating effective coverage cascades

#### Target population (step 1)

The target population is both the first step of the cascade and the denominator for every other step of the cascade; thus, obtaining an unbiased estimate is critical. We used population-based data (household surveys) to identify the target population in order to minimize selection bias. The target population was defined as women with a live birth in the two years prior to the survey (for ANC and PNC cascades) or children under five years with symptoms of illness that should prompt care-seeking or treatment (fever, diarrhea, and suspected acute respiratory illness).

For small and/or sick newborn care, we also used women with a live birth in the two years prior to the survey as the target population. This is because DHS and MICS do not identify small or sick newborns. Although DHS includes questions that collect birth size, these questions do not provide valid measures of LBW [18] [19]. Thus, for small and/or sick newborn care, we had to use an alternative approach. We considered that all health facilities providing childbirth services need some capacity to manage small and/or sick newborns, whether to provide definitive care or to stabilize and refer to a higher-level facility. The target population for our analysis was therefore live births in the two years prior to the survey.

#### Service contact coverage (step 2)

Service contact coverage is the proportion of the target population who visits a qualified health service or is visited by a non-facility provider (**Table 1**). We calculated service contact coverage from household survey data, using standard indicators and definitions for service contact coverage indicators [20,21], including ANC coverage, institutional delivery, PNC coverage, and care-seeking for children with fever, difficult breathing, or diarrhea.

We limited the service contact coverage numerator to women and children who had sought care from a “qualified” health facility or provider. “Qualified” refers to country policy on service delivery rather than a specific facility’s readiness or provider competence. For example, in some countries community health workers (CHWs) are permitted to provide care for sick children; in countries with this policy, sick children taken to a CHW would be included in the numerator of the care-seeking indicator, as would children taken to a public, private, or faith-based health facility. We did not consider pharmacies to be qualified providers, as they typically do not formally assess or diagnose patients.

#### Readiness and process quality-adjusted coverage (steps 3 and 5)

##### Measures of service readiness and process quality

For EC cascade steps 3 (input-adjusted coverage) and 5 (process quality-adjusted coverage), we needed summary measures of the readiness of health facilities to offer services and interventions, and the quality with which these services and interventions are provided. We used health facility survey data from existing SPAs and SARAs to estimate facility readiness and process quality (**Table 1**).

There is no global consensus or guidance as to how readiness and process quality should be estimated for services along the RMNCAH&N continuum, and as a result, approaches and measures have proliferated [9]. In most cases, a facility requires multiple items to be “ready” to deliver an intervention, or to deliver the intervention with high quality. For health services that serve as platforms for the delivery of multiple interventions, a very large number of items may be collected to evaluate health facility readiness and process quality; for example, studies that have identified relevant facility readiness and process quality items through a mapping of service guidelines found 121 relevant items for ANC [22] and 866 relevant items for small and/or sick newborn care [23]. Multiple approaches have been used to generate readiness and process quality indices from these items, including data reduction techniques such as principal components analysis, simple or weighted averages, and use of expert opinion and clinical guidelines to prioritize the most relevant items [9].

We used a four stage approach to develop readiness and process quality measures for our service areas of interest, wherein we: (1) identified globally recommended interventions, based on WHO and UNICEF guidelines; (2) extracted facility readiness and provision of care items from intervention-specific clinical and service implementation guidelines; (3) mapped the identified items from the guidance documents to available data in health facility surveys; and (4) developed indices informed by quality of care frameworks, clinical guidelines, and data availability [22,24,25]. For some service areas where a very large number of items were identified (ANC, sick child care, maternal and child nutrition), we also conducted an expert survey to prioritize the items to be included in the indices. Depending on the service area, we used simple averages or domain-weighted averages to calculate summary measures, where domains were either interventions or categories of items (e.g., infrastructure, human resources, drugs, diagnostics). All scores were scaled from 0 to 1, with a score of 1 indicating perfect readiness or process quality and 0 indicating no readiness or process quality. Detailed descriptions of the development of these measures have been published separately [22,24,26].

Although we developed readiness and service provision indices for our analyses, we do not recommend the use of a particular index or approach. However, Box 1 highlights some factors to consider when developing these indices.

###### Box 1. Factors to consider in developing readiness and process quality indices for effective coverage cascades

1. **How will you decide which items to include in the index?** Decisions about which items to include can be based on conceptual frameworks, clinical guidelines, expert opinion, and/or data reduction techniques. Framework- and guideline-driven approaches may be easier to explain to stakeholders, more replicable, and have greater face validity, as it is possible to ensure the inclusion of key items. Data-driven approaches like principal component analysis make use of the data itself to identify the elements that explain most of the variation in readiness or quality but may be more difficult to explain and interpret; may not explain a large fraction of the variation; and can lead to counterintuitive results.
2. **Will your indices be adapted to country guidelines and practices, or standardized across countries?** Indices that are primarily intended to estimate readiness- or quality-adjusted coverage for a specific country should be tailored to country guidelines and practices, by or in collaboration with the Ministry of Health and local researchers. For multi-country analyses, however, it may be necessary to define a standard set of items that can be included for all countries – this may result in the loss of some country specificities.

##### Linking household and health facility data

To estimate readiness-adjusted (step 3) and process quality-adjusted (step 5) coverage, we linked the summary measures of readiness and process quality calculated from the HFA to household survey data. This process required assigning a readiness or process quality score to each care-seeking episode in the household survey dataset. There are two potential ways to implement this linking: assigning the process quality or readiness score for the specific facility that the woman/child visited during the care-seeking episode (exact-match linking); or assigning an average process quality or readiness score to the care-seeking episode (ecological linking) [27]. Exact-match linking requires information on the names of health facilities, pharmacies, and/or CHWs visited. This information is usually not collected in household surveys, and facility names are typically not available in health facility assessment (HFA) datasets. Three studies have compared ecological linking to exact match linking and have found them to be generally equivalent when the ecological linking method accounts for the type of facility visited by the woman or child [28–30].

We used ecological linking by stratum, where care-seeking episodes were linked to a stratum average readiness or process quality score. Strata were defined by the types of facility visited (facility level and managing authority, where available), and by the sub-national administrative area where the woman or child resided (for example, region or district). In each country there were differences between the health facility categories in the two data sources. HFA facility categories tended to be more detailed, while health facility categories in the household survey were broader, reflecting what survey respondents might be able to report. We mapped household survey facility categories to HFA categories, collapsing the HFA categories where needed such that every household survey facility category mapped to a single HFA facility category. We then calculated mean readiness and process quality scores for each stratum. These stratum mean scores were merged with the household survey data by care-seeking facility category and administrative area, so that each care-seeking episode from a qualified provider had a readiness (and process quality, if available) score assigned to it. Thus, for example, records for women in Sylhet division, Bangladesh, who reported attending ANC at an Upazila Health Complex (UHC), were assigned a mean ANC readiness score calculated across UHC facilities in Sylhet. Women or children for whom care was not sought from a qualified provider were assigned a score of 0. We then calculated the mean readiness or process quality score across the target population in the household survey dataset, yielding the readiness-adjusted or process quality-adjusted coverage.

In some countries, there were empty strata in the HFA dataset, meaning that not all facility types were sampled in each administrative area. As a result, some care-seeking episodes in the household survey dataset could not be linked to an average readiness or process quality score using strata defined by facility type and administrative area. This was particularly an issue in settings with many facility types and/or many administrative units, resulting in very fine strata (for example, Tanzania had over 350 strata because of the large number of facility types and regions). For the ANC cascades, the proportion of empty strata ranged from 0% in Bangladesh and Nepal to 50% in Tanzania. In cases where empty facility strata prevented linking by facility type and administrative area, we used a less granular approach to linking. We re-defined facility strata by facility category, ignoring administrative area, and used these averages to link to household survey data.

##### Care-seeking from multiple sources

For certain service contact coverage indicators, the household survey respondent can report care-seeking from multiple sources. For our analysis, this was the case for ANC and for sick child care. In these cases, we calculated the mean readiness and process quality scores across all qualified facility types reported by the survey respondent.

##### Multiple observations per facility

Measures of process quality are often based on multiple individual observations of care. The number of observations conducted at a facility for a SPA depends on the number of clients who seek care on the day of the assessment as well as the workload of the data collection team [14]. When linking HFA data to a household survey, there are two possible approaches to analyzing these data: first taking the mean of the observations for each facility to obtain a mean facility process quality score, and then calculating a mean process quality score across the facilities in each each stratum; or second, calculating the mean across all observations within a stratum, without accounting for facility. We chose the second option so that the stratum averages would be representative of patient care overall rather than representative of health facilities. In addition, this approach implicitly weights health facilities by the number of observations conducted in the facility. We considered this to be an advantage because we expected the number of observations to be correlated with facility caseload [31], and previous work has found that weighting readiness and provision of care scores by caseload improved the accuracy of ecological linking [29].

#### Intervention coverage (step 4)

We calculated intervention coverage estimates from DHS or MICS data (**Table 1**). For each service area, we attempted to identify relevant biomedical, counselling, or behavioral interventions measured in the household survey. The number of interventions included varied by service area, ranging from none (small and/or sick newborn care) to seven (ANC). We used standard intervention coverage definitions [20,21] and calculated the mean intervention coverage value for each service area.

We calculated intervention coverage among the entire target population. We did not limit our intervention coverage measures to women or children who reported care-seeking from a “ready” health facility, because readiness was not measured contemporaneously with reported care-seeking. Additionally, we could not match survey respondents to the specific facility that they visited, and it is possible for a facility to have a moderate or even low readiness score and still deliver certain interventions. We also did not limit intervention coverage to women or children who sought care from a qualified health facility or provider, as some interventions can be delivered appropriately by other sources. For example, pregnant women may obtain iron folic acid (IFA) supplements from pharmacies, and caregivers may purchase oral rehydration solution (ORS) from corner shops.

#### User adherence-adjusted coverage (step 6)

We did not estimate user adherence-adjusted coverage for any service area because there were few or no valid measures of adherence in DHS or MICS for our service areas of interest. For example, for ANC, the only measure of adherence in DHS is consumption of iron-containing supplements, and there is evidence that this question may be difficult for women to answer accurately [32,33]. For sick child care, DHS and MICS do not typically collect information about adherence.

#### Outcome-adjusted coverage (step 7)

We also found several challenges in measuring outcome-adjusted coverage. First, as noted by Marsh et al, service packages deliver many interventions with different outcomes, making it difficult to identify a single health outcome [6]. This is particularly the case for preventive services, such as ANC and PNC. In addition, the health or nutrition outcome should be attributable to the intervention or service area [5,6]. Many of the outcomes measured in DHS and MICS (all-cause neonatal mortality; all-cause under-five mortality; acute and chronic malnutrition) are relatively broad and influenced by many factors, making attribution challenging, particularly in a cross-sectional survey.

#### Weights

Service contact coverage (step 2) and intervention coverage (step 4) were weighted using the DHS women’s weights or the MICS child weights. For input-adjusted (step 3) and process quality-adjusted (step 5) intervention coverage estimates combine household and facility data, we applied multiple weights [20,34]. For input-adjusted coverage (step 3), we first weighted stratum mean readiness scores by the facility weights provided in the SPA and SARA. These weights combine sampling and non-response weights for sampled facilities. After linking the readiness data to the household survey data, we then weighted the input-adjusted coverage estimates using the DHS women’s sampling weights, or, in the case of the MICS, the child sampling weights. For process quality-adjusted coverage (step 5), we followed the same process as for readiness-adjusted coverage, except that the stratum mean process quality scores were weighted using SPA client weights, which combine the facility sampling weight, the client sampling weight, and a non-response weight.

#### Variance estimation

We used a design-based approach [20,35,36] to estimate variance of household survey measures, i.e., service contact coverage (step 2) and intervention coverage (step 4). Taylor linearization was used to account for the effects of cluster sampling [37].

Variance estimation for readiness- and process quality-adjusted coverage (steps 3 and 5) was more complex. When coverage estimates are constructed by combining data from different sources, each source is subject to sampling error, such that all sources of variation contribute to the estimate’s precision. Even in the case of facility censuses, where all health facilities are surveyed, health workers and consultations are still subject to random selection processes, which generates a sampling error. Recent studies have suggested the delta method to estimate the total variance of readiness- or process quality-adjusted coverage estimates using asymptotic properties of coverage estimators [38]. These methods make several assumptions about the statistical properties of coverage estimates: first, there is an assumption about the centrality of the distribution for both traditional coverage estimates as well as the estimates derived from facility surveys, and second, that samples are large enough for asymptotic behavior to apply to effective coverage estimators [39]. The delta method, while straightforward to use, is also difficult to adapt to specific features of an EC analysis, such as linking by geographic areas or accounting for multiple sources of care.

An ideal method for estimating the total variance of readiness- or process quality-adjusted coverage estimates would account for variability from all sources and also allow for flexibility in the linking method, while providing good coverage at relatively small sample sizes, which are not uncommon in HFAs. While parametric methods including the delta method and the parametric bootstrap related to EC estimates have been examined previously by Sauer et al [38], to our knowledge non-parametric methods have not been examined. Jackknife estimators for multistage surveys have well-established statistical properties and are widely used in the analysis of complex surveys [40,41]. A jackknife estimate of the standard error has several properties likely to be advantageous, including limited assumptions regarding the distribution of sample survey estimates [42], suggesting that the jackknife would be a robust estimator. The jackknife estimator is also more flexible than parametric methods, making for more involved implementation, but also allowing for adaptation to analytical methods. For our analysis, we used a jackknife approach to estimate the standard errors for readiness- and process quality-adjusted coverage, where the standard error was derived from the distribution generated by withholding each household cluster and each health facility.

### Challenges in estimating effective coverage cascades for RMNCAH&N

#### Feasibility

Using available DHS/MICs and SPA/ SARA data, we were able to estimate EC cascades through intervention coverage (step 4) for one service area (maternal PNC and newborn PNC), and through process quality-adjusted coverage (step 5) for four service areas (ANC, sick child, maternal and child nutrition) (**Table 3**). No process quality data on PNC were collected in SARA or SPA, so we were unable to estimate process quality-adjusted coverage for PNC for mothers or babies. For one service area, small and/or sick newborn care, we did not have an appropriate denominator or measure of service contact coverage (**Table 3**). Although we still attempted to estimate a cascade for small and/or sick newborn care, we made substantial adjustments, as described above, using all live births as the target population and institutional delivery as the service contact coverage measure. In addition, there were substantial gaps in the readiness data available for small and/or sick newborn care. The resulting cascade was potentially limited in its utility.

**Table 3.**
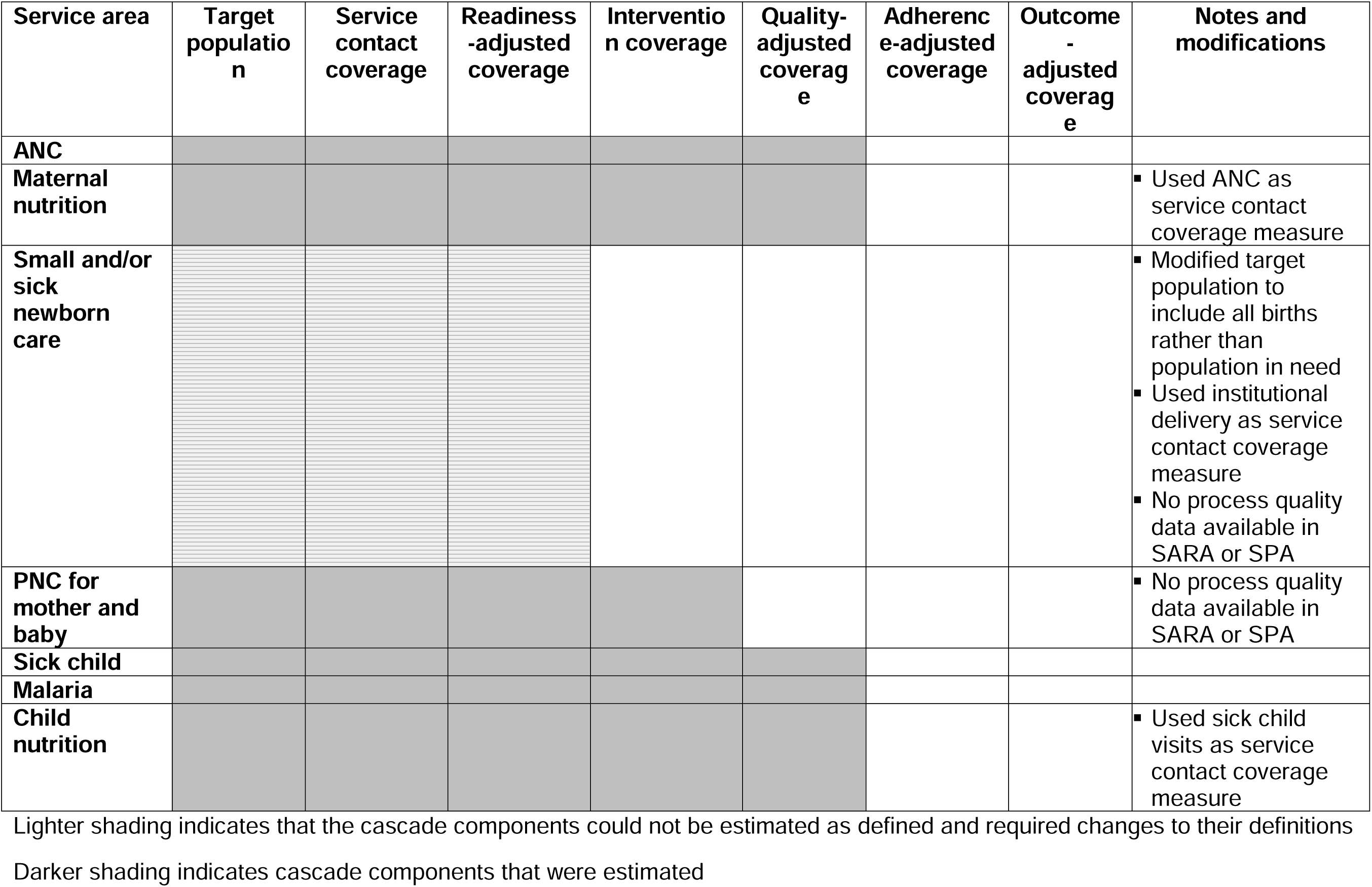
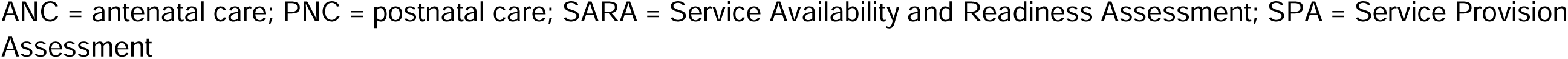
Feasibility of effective coverage cascade estimation using survey data, by service area*.

Although we estimated coverage cascades for maternal and child nutrition, there were no service contact coverage measures in the household survey on care-seeking for nutrition services (e.g., growth monitoring, treatment of acute malnutrition) or for nutrition services delivered outside the health facility (**Table 3**). As a result, we focused on nutrition interventions delivered through other service contacts: ANC and sick child visits. However, these represent a small sub-set of nutrition interventions for women and children. We were not able to estimate readiness-adjusted or process quality-adjusted coverage (steps 3 and 5) for nutrition interventions outside of these service contacts.

We were not able to estimate adherence- or outcome-adjusted coverage (steps 6 and 7) for any service area. As described above, these steps of the cascade are challenging to measure for service areas that include many interventions, and this is exacerbated by limited data on adherence and RMNCAH&N outcomes in existing population-based surveys. Marsh et al and Amouzou et al also noted these challenges and Marsh et al suggested using process quality-adjusted coverage as a proxy for outcome-adjusted coverage in cases where outcome-adjusted coverage is not feasible or appropriate to measure, while Amouzou et al. suggested that cohort studies might be a useful approach to measuring outcome-adjusted coverage [5,6].

#### Data limitations

Data gaps and limitations were the largest barrier to the estimation of EC cascades for RMNCAH&N in our analysis. The most commonly available and accessible data were national household surveys, which provided information on the target population, service contact coverage, and intervention coverage. Household surveys are conducted approximately every three to five years in many low-income countries and some middle-income countries; however, some had gaps of seven to ten years or more between household surveys.

Existing household surveys also had some content gaps: they did not measure service contact coverage for small and/or sick newborns, for treatment of acute malnutrition, or for well-child visits at which children might receive preventive interventions. In addition, household surveys collected limited information on the interventions delivered during certain types of services, largely because of the difficulty of measuring these interventions accurately in a household survey (see, for example [43–47]).

In contrast to household surveys, HFAs had large geographical gaps as well as less frequent surveys. We identified 39 countries that had conducted a SPA or SARA/HHFA in the ten-year period from 2009 to 2018 (five that had both a SPA and SARA/HHFA, five only SPA, 29 only SARA/HHFA); of these, ten countries had publicly available datasets.

Health facility surveys also had content gaps around readiness and process quality data. Service area-specific gaps in HFAs have been described in previous publications mapping the availability of readiness and process quality items [22,24]. In addition, the lack of data on provider competency was a major cross-cutting gap. The presence of a skilled provider is essential to providing high quality care, but most HFAs do not measure provider competency and instead collect data on whether providers received training in a particular area in the previous two years as a proxy.

#### Estimating effective coverage of community- and home-based interventions

The EC cascade framework includes service contact coverage at its foundation, and thus by definition is focused on interventions delivered through health services, primarily at health facilities. This is further reinforced by HFA sampling frames, which generally exclude community-based workers. Community- and home-based interventions are also not currently captured in some countries’ health information systems. However, essential RMNCH&N community- and home-based interventions (e.g., community case management of childhood illness and malnutrition, and counselling on infant and young child feeding practices), as well as interventions delivered outside the health sector, such as for water, sanitation, and hygiene (WASH) interventions, are recommended by WHO.

For our analyses, we categorized CHWs as qualified providers in settings where policy allowed the CHWs to provide the interventions in questions. For the readiness- and process quality-adjusted analyses, because we had no data on CHWs, we assumed that their readiness and process quality was the same as for public first level facilities. We also did not restrict our intervention coverage measures to women or children who had received care in a health facility. However, EC cascade frameworks and methods may need further adaptation to reflect the effective coverage of community- and home-based interventions. For community-based care, this may include counting CHWs as qualified providers of a health service and seeking data on their readiness and process quality. For home-based and non-health sector interventions, the cascade itself may need to be modified.

#### Nested estimates

The coverage cascades proposed by Tanahashi, Amouzou, and Marsh [5,6,8] are nested, meaning that an individual’s inclusion in the numerator of one step of the cascade is conditional on their inclusion in the numerator of the previous step. For example, Step 4: intervention coverage – the proportion of the target population receiving an intervention – is measured only among the proportion that sought care from a ready facility (Step 3: readiness-adjusted coverage). This presented analytical challenges.

Nesting poses analytical issues when the data for different steps of the cascade come from different data sources, source populations, and/or timepoints. In these cases, the data in actuality are not nested. For example, in the ANC EC cascade, we observed that mean intervention coverage (Step 4) was greater than readiness-adjusted coverage (Step 3) for 2 countries for the first visit (ANC1) cascade (Nepal and Sierra Leone) and for all countries for the four visit (ANC4) cascade. This indicates that some women who attended ANC from a facility type with low average readiness nonetheless reported receiving the coverage interventions measured in DHS (e.g., IFA, tetanus toxoid vaccine, IPTp, deworming, blood pressure measurement, blood draw, urine sample). There are at least two possible reasons for this. First, we used ecological linking, meaning that we linked women to an average facility readiness score, rather than the readiness score for the facility the woman attended. Thus, it is possible that some women attended a high readiness facility but were assigned a low readiness score because, on average, facilities of that type in the woman’s region had low readiness. Second, HFAs measure readiness and process quality at a specific point in time, whereas in our ANC analysis intervention coverage was measured over a two-year recall period. Thus, the readiness and process quality at the time of the HFA may not reflect the readiness and process quality at the time a woman received services at the facility. In cases like these, analysts must decide whether to calculate intervention coverage directly, resulting in a cascade that can decrease or increase from step to step, or whether to scale intervention coverage by facility readiness, which would result in a nested cascade but might under-estimate true intervention coverage.

#### Cascade interpretation

EC cascades provide more information than a single coverage, quality, or outcome indicator: the set of steps along the cascade, taken together, provide information about the reasons why a population may not fully benefit from a health service. Interpretation of a RMNCAH&N EC cascade focuses on the gaps between the steps of the cascade (access gaps, readiness gaps, process quality gaps), which can identify areas for prioritization in national health and nutrition plans or by funders [5,6]. However, the EC cascade alone does not provide detailed information about how to address these gaps, i.e., the reasons for low service contact, readiness, or process quality. In addition, while the summary readiness and process quality indices provide an overall picture of readiness or service quality the indices did not provide information on specific interventions. Thus, in many situations, it may be important to also present the detailed service access, readiness, and process quality data that underlies the estimates in an EC cascade. To explore ways to address this issue, in each of the service area-specific EC cascade publications, we included information on the sources of care; availability of specific readiness and process quality items; domain-specific readiness and process quality scores; and the distribution of readiness and process quality scores by facility type and managing authority. Much more work is needed to assess the utility of this type of information for decision-making at global, national, and sub-national levels.

## Discussion and recommendations

This paper presents a description of the methods used to estimate EC cascades for RMNCH&N using existing, publicly available survey data and a discussion of the methodological choices and challenges identified. This work adds to a body of literature that has estimated EC using various methods, particularly over the past decade [7,9,48–53]. Several reviews [5,9,27] of studies reporting EC or quality-adjusted coverage have noted variability in the definitions and methods used. To address this variability, in 2020, Marsh et al published definitions for EC and for the components of EC cascades [6]. This paper presents a detailed description and justification of the methods we used and the issues we encountered when trying to operationalize the EC cascades proposed by Amouzou et al and Marsh et al. Where possible, we have based our methodological choices on empirical data, but we note methodological gaps and the need for further research in certain areas.

Some of the analyses to estimate EC cascades were relatively straightforward: we used standard methods and indicators to estimate service contact coverage and intervention coverage using household survey data. However, in order to estimate readiness- and process quality-adjusted coverage, we had to first define summary measures of readiness and process quality for each service area, and then “link”, or combine, household survey data with these summary measures. There was very little guidance on how to define and calculate summary measures of readiness and process quality, although there was a large diversity of published approaches. As a result, we developed our own summary measures and documented the process [22,24,26], but we emphasize that these are illustrative and not intended to be definitive summary measures for these service areas. Instead, countries should adapt existing summary measures or develop their own service area-specific summary measures based on the data that is available to them and their health plans and priorities. We also found that even standardized HFAs like the SPA and SARA varied from country to country and over time, perhaps in order to reflect national priorities, and this may make it challenging to define a single readiness and process quality index that would be comparable between countries and surveys.

In contrast to the development of summary measures of process quality and readiness, the process for linking household and health facility data was relatively straightforward and was supported by a body of empirical methods work [28–30,54–56]. However, we found that methodological decisions or development were still necessary regarding variance estimation, weighting health facility data [31], accounting for multiple sources of care, and aggregation of client visit data. We also noted four main issues that arose during cascade development: first, data gaps; second, challenges in estimating EC of home- and community-based interventions; third, challenges in nesting estimates along the cascade; and fourth, challenges in ensuring that the results were interpretable and actionable.

Like other authors, we found that data availability was an important barrier to the estimation of the proposed EC cascades [5–7,17]. We found that data gaps were more severe for readiness and process quality data than for service contact coverage and intervention coverage. One limitation is that we used only publicly available data from HFA; data from routine health information systems has been proposed as an alternative source that may fill these gaps [57]. However, recent reviews of RHIS for RMNCAH&N also found substantial gaps and variability across countries, as well as accessibility issues [17,58–60]. For EC cascades to be used for decision-making, more frequent measures of readiness and process quality will be needed.

The challenges in estimating EC cascades for home- and community-based interventions have been raised previously [28,54]. These delivery modes are critical to many RMNCAH&N service areas, particularly for nutrition, child health, and WASH interventions, hence our suggestion to develop EC cascades for other service delivery models.

To our knowledge the challenges in nesting the EC cascades have not been previously mentioned. Although Marsh et al noted the need to bring together data from multiple sources to estimate the full cascade [6], we found that using multiple data sources made it difficult to estimate a true cascade with nested estimates. The difficulty in nesting data from different sources may make it more attractive to use EC cascade approaches based on single data sources rather than linked sources. As noted above, work has begun on using RHIS data to estimate EC, and much more work in this area is needed. Similarly, Arroyave et al have proposed an ANC process quality measure based on household survey data [61], which could allow much of the ANC cascade to be estimated from household survey data. Because of limitations in what survey respondents can report, this approach will not be feasible for all service areas, but should be considered for certain services, including community- and home-based interventions.

Our analyses had a number of limitations. First, we relied on publicly available data, which in practice meant that we used DHS, MICS, SPA, and SARA data. It is possible that non-publicly available datasets might have addressed some of the data gaps that we have noted, particularly around provision of care. Because these analyses were primarily an analytical and methods development exercise, we also did not work with countries to adapt or validate the cascades. For cascades that are intended to be used in and by individual countries, the EC cascade analysis, including the development of the theoretical cascade, identification of data sources, development of summary quality measures, and analysis and interpretation of data, should be done at country-level and in close collaboration with researchers and Ministry of Health officials.

Our analysis also focused on service areas rather than individual interventions. Although this approach provides a broad assessment of the potential health gain from key RMNCAH&N services, it can make it more challenging to estimate adherence-adjusted and outcome-adjusted coverage, as many interventions within a service area have no adherence component, and it can be difficult to identify a measurable, strongly associated outcome for an entire service area. Our study also did not include empirical work on the interpretation and usability of EC cascades at global, national, and sub-national levels, but previous work has noted this as a key research priority [6,9].

EC is one approach to identifying and quantifying bottlenecks to improving RMNCAH&N outcomes. We found that it was feasible to estimate EC cascades for many RMNCAH&N service areas using existing, publicly available data, and we have described our methods and experience to support research and practice in this field. However, we have also noted several areas where additional research and development is needed, which we have summarized in Box 2. In order to transition EC cascades from research to practice, it will be essential to provide clear guidance on methods and definitions, account for the full spectrum of RMNCAH&N interventions, and understand how EC cascades can be interpreted and used by stakeholders to improve health systems and programs.

### Box 2. Recommendations for effective coverage cascade research and practice for RMNCAH&N

1. In addition to the foundational work of Amouzou et al and Marsh et al [5,6], there is a need for **consensus on analytical methods for estimating RMNCAH&N EC cascades and interpretation of the cascades**, based on experiences estimating and using RMNCAH&N EC cascades and their components for different services and in different settings.
2. There is currently no **consensus guidance on summary measures of readiness and process quality for RMNCAH&N services**; the development of evidence-based guidance would facilitate the estimation of EC cascades.
3. Geographic and content related data gaps have limited our ability to estimate RMNCAH&N EC cascades. A major priority is **increasing the availability of RMNCAH&N quality of care data, including readiness and process quality data**.
4. There is a need for research **on EC cascades, methods, and data sources for non-facility-based RMNCAH&N services and interventions**.
5. Applied research is needed at national and sub-national levels to **identify best practices for adapting RMNCAH&N EC cascades for health services and for individual interventions, presenting the cascades to stakeholders, and using the results for planning and decision-making to drive policy-making and programmatic change**.

## Data Availability

DHS and SPA data are publicly available through the DHS program: https://dhsprogram.com and MICS data are available through Unicef at: https://mics.unicef.org/. We obtained permission from the Sierra Leone Ministry of Health to use the de-identified 2017 SARA data.

https://dhsprogram.com

## DECLARATIONS

### Acknowledgements

The authors wish to acknowledge the Bill & Melinda Gates Foundation for their support of this project. We are grateful to the Improving Measurement & Program Design (IMPROVE) Coverage Group for their technical inputs and expertise during the development of this manuscript. In addition to the co-authors, the IMPROVE Coverage Group includes:

- Agbessi Amouzou, Johns Hopkins University
- Fred Arnold, ICF International (retired)
- Ann Blanc, Population Council (retired)
- Harry Campbell, University of Edinburgh
- Louise Tina Day, London School of Hygiene and Tropical Medicine
- Thom Eisele, Tulane School of Public Health
- Attila Hancioglu, MICS (retired)
- Joanne Katz, Johns Hopkins University
- Sunny Kim, International Food Policy Research Institute
- Margaret Kosek, The University of Virginia
- Tanya Marchant, London School of Hygiene and Tropical Medicine
- Allisyn Moran, World Health Organization
- Moise Muzigaba, World Health Organization
- Jennifer Requejo, United Nations International Children’s Emergency Fund

### Ethics statement

This is a secondary analysis and as such did not involve human subjects research.

### Data availability

DHS and SPA data are publicly available through the DHS program: https://dhsprogram.com/Data/ We obtained permission from the Sierra Leone Ministry of Health to use the 2017 SARA data.

### Funding

This work was supported by the Improving Measurement and Program Design grant (OPP1172551) from the Bill & Melinda Gates Foundation. The funding agency had no role in the design of the study, analysis, interpretation of data, or writing of the manuscript.

### Authorship contributions

MKM, AS, and EC conceptualized the paper; MKM, AS, EC, and JP conceptualized the analyses; AS and EC conducted the analyses; MKM wrote the manuscript, and all authors critically reviewed and revised the manuscript.

### Disclosure of interest

The authors completed the ICMJE Disclosure of Interest Form (available upon request from the corresponding author) and all disclose no relevant interests.

## Notes

### Competing Interest Statement

The authors have declared no competing interest.

### Author Declarations

This study was a secondary analysis of survey data available from https://dhsprogram.com/ and https://mics.unicef.org/

